# Has the pandemic enhanced and sustained digital health-seeking behaviour? A big data interrupted time-series analysis of Google Trends

**DOI:** 10.1101/2022.07.29.22278191

**Authors:** Robin van Kessel, Ilias Kyriopoulos, Brian Li Han Wong, Elias Mossialos

## Abstract

**Background:** Due to the emergency responses early in the pandemic, the use of digital health in healthcare increased abruptly, yet it remains unclear whether this introduction was sustainable on the long term. We explore trends in digital health-seeking behaviour as proxy for readiness to adopt digital health as a mainstream form of healthcare.

**Methods:** We use weekly Google Trends data from February 2019 to August 2021 in Canada, United States, United Kingdom, New Zealand, Australia, and Ireland. We used five keywords to monitor online search interests in Google Trends: *online doctor, telehealth, online health, telemedicine*, and *health app*. Data are analysed using an interrupted time-series analysis with break-points on 11 March 2020 and 20 December 2020.

**Results:** Digital health searches immediately increased in all countries after the pandemic announcement. There was some variance in what keywords were used per country. However, searches declined after this immediate spike, sometimes towards pre-pandemic levels. The exception is the search volume of *health app*, which showed to either remain stable or gradually increase during the pandemic.

**Interpretation:** Our findings suggest that digital health-seeking behavioural patterns associated with the pandemic are currently not sustainable. Further building of digital health capacity and development of robust digital governance and literacy frameworks remain crucial to more structurally facilitate digital health transformation across countries.

**Funding:** Not applicable.

## Introduction

On 11 March 2020, the World Health Organization formally announced that COVID-19 was classified as a pandemic, which disrupted healthcare services worldwide. Evidence from a systematic review shows a median 42% decrease in healthcare visits, 28% decrease in admissions, 31% reduction in diagnostics, and 29% reduction in therapeutics, resulting in an overall 37% decrease of utilisation of healthcare services (1). Digital health usage has exploded during the early stages of the COVID-19 pandemic as traditional health services were disrupted (2–4). Digital health generally refers to the use of internet solutions, big data, and communications technologies to collect, share and manage health information to improve both individual and public health, as well as identify symptoms, plan treatment, monitor key health parameters, and monitor progress and treatment effects (5,6). Even though digital health saw tremendous uptake early in the pandemic (4), no information exists on whether this uptake was sustainable (7), which we investigate in this article.

Due to the sudden disruption of traditional health services, health ecosystems may have digitalised predominantly out of a need to survive rather than a desire to innovate, highlighted by the emergency transition to a digital paradigm and subsequent changes when the pandemic started (2). Implementing digital health solutions is complex and relies on many institutional factors, cultural and behavioural traits and health system characteristics (7,8). For instance, the design process of digital health solutions is indicative of which populations it will be able to reach and, equally importantly, what population groups will experience difficulties in accessing and using the tool (3,9). Policy environments are also vital to laying the foundation of how conducive a health system is to adopting a digital health solution and how health professionals are trained in the field of digital health (3,9,10). Finally, the readiness and willingness of digital health users are crucial elements in adopting digital health solutions (2,8). A recent analysis of digital skills in the European Union – frequently regarded as a leading region in terms of digital skills – found clear discrepancies across the European region (11). Certain population groups also seem to fare more favourable in a digital world: people who are younger, higher educated, male, live in urban regions, and are either a student or employed consistently report higher internet access and digital skills (11– 14). These findings suggest that − if digital health services were structurally introduced now − only certain population groups could fully benefit from these services. Paradoxically, population groups that have higher needs for healthcare and could potentially benefit most from these innovations are the ones that would experience the highest barriers to access (9).

With traditional health services having regained their ability to function, patients can increasingly decide whether they prefer to seek traditional or digital healthcare, which often starts with online search engines (15). Although the internet cannot substitute health professionals as a sole health information source due to a combination of the prominence of misinformation and a lack of health, digital, and science literacy (13,16–18), search engine data can be instrumental in understanding the general preference of populations in exploring the possibility of using digital health (19,20). While online searches can be information- or curiosity-driven, previous research has shown that online search behaviour is strongly correlated with the actual (healthcare) needs of the population and forms an integral part of a pathway to actual communication with healthcare providers (19–22).

This article aims to further explore the digital readiness for structurally implementing digital health solutions by investigating the health-seeking behaviour of the general population by analysing Google Trends™ data. These data showcase how popular specific search terms were in certain countries. The main advantage of Google Trends™ is that it uses the revealed and not stated users’ preferences (23), making it possible to obtain data that would be difficult to collect otherwise. The advantage of revealed preference data is that it is based on actual decisions, meaning there is no need to assume that participants will respond to simulated situations as is the case with stated preference data. As a result, revealed preference data are characterised by high reliability and face validity (24). Google Trends™ have shown to be a viable tool to understand, monitor, and even forecast information-seeking trends and public interest and is becoming an increasingly popular method for assessing population preferences in health research (19,25,26).

## Methods

Google Trends™ is the principal tool used to study trends and patterns of search engine queries using Google (19). It is an openly accessible big data tool that provides real-time and archived information on Google queries from 2004 onwards (23). Data are available in realtime, solving issues that arise with conventional, time-consuming survey methods (23). So far, it has been applied to various health-related topics such as mental health, vaccine hesitancy, and infodemic surveillance (22,27,28).

All Google Trends data points are normalised and scaled (23), meaning the number of searches for a specific term is divided by the total number of searches for all topics at a particular location and within the specified timeframe, resulting in a normalised score. All normalised scores are scaled to have a maximum score of 100 points. Scaled scores (known as relative search volumes) range from 0 to 100 points and represent search interest relative to the month with the highest search interest. Google adjusts relative search volumes for internet access and population size. We followed the established methodological guidelines for using Google Trends in infodemiology and infoveillance (23).

### Keyword, Timeframe, and Country Selection

Based on the keywords used in recent systematic reviews in the field of digital health (29– 31), we used five keywords to monitor online search interests in Google Trends: *online doctor, telehealth, online health, telemedicine*, and *health app*. While we also explored *digital health, digital therapeutics, telecare, telemonitoring*, and *virtual health*, these terms’ search volumes were negligible (< 1). As such, we did not include them in our analyses. We extracted weekly relative search volume data for Canada, the United States, the United Kingdom, New Zealand, Australia, and Ireland from 1 February 2019 to 1 August 2021 (n=780 country-weeks). The six countries were chosen because they share English as their dominant language and provide a varied representation in policy landscape regarding digital health (9,10). Google is used for 87% to 93% of the online search queries in the countries under study (32–34), meaning our data accurately captures the preferences of the population of the countries under study. As such, we can accurately measure public interest in digital health in the midst of a global pandemic (25).

### Empirical approach

To assess both short- and long-term effects of the pandemic on digital health search behaviour, we used a single-group interrupted time-series design (35,36), which is suitable for evaluating population-level effects of critical events (e.g., interventions, policy innovations, or public health crises) that have occurred at a clearly defined point in time (37).

Our design adopts a segmented regression analysis approach that allows for the identification of three effects by comparing pre- and post-event trends (37,38): (i) the pre-event slope, indicating how the outcome of interest was changing prior to the occurrence of the critical event; (ii) the change in intercept, identifying the immediate change following the critical event; and (iii) the post-event slope, capturing the gradual change in the period after the critical event. In this article, the critical events are the start of the pandemic on 11 March 2020 and the announcement of the vaccines on 20 December 2020. The start of the pandemic was chosen as a critical event because of the shock it brought to health system functioning and the public attention it raised about the health risks associated with COVID-19 (1). The vaccine announcement was chosen as a second critical event as this signalled the start of a new period where it was possible to partially return to the pre-pandemic status quo (39).

This approach addresses internal validity constraints and represents a methodologically robust design for measuring the impact of critical events (40), and has been widely used in empirical work in the field of public health (41–43). We also computed seven-day moving averages for active COVID-19 cases and COVID-19 deaths using Oxford Government Policy Tracker data (44). These moving averages were added as covariates in the segmented regression model, as trends in COVID-19 cases and deaths may be associated with (digital) healthcare-seeking behaviour during the pandemic. Our segmented regression models were fit using Newey-West standard errors, which are designed to account for autocorrelation and potential heteroskedasticity in time-series data (45). We conducted a post-estimation analysis to capture the exact post-intervention trend.

To ensure that we fit a model that accounts for the correct autocorrelation structure, we performed Cumby-Huizinga tests for autocorrelation for each individual segmented regression model (46). The appropriate lag was determined by performing Cumby-Huizinga tests on segmented regression models without lag. Finally, we conducted a sensitivity analysis to verify the robustness of the segmented regression model, consistent with a placebo intervention using the time period 1 February 2017 to 1 August 2019. Data were extracted using R version 4.1.2 and the analysis was performed using STATA version 17.0.

## Results

### Descriptive analysis

When comparing raw data searches before and after the pandemic announcement on 11 March 2020 (shown in Figure 1), we observe an immediate increase in the relative search volumes of *online doctor, online health, telehealth*, and *telemedicine*. The search volume of *health app* did not indicate an immediate reaction, though the search volumes increased over time. After the immediate increase, we also observed a gradual reduction in the search trends of *online doctor, online health, telehealth*, and *telemedicine*, while *health app* remained stable. Furthermore, there is limited reason to assume “anticipation” as a relevant factor in digital healthcare-related search behaviour, given that search volume substantially increased only after the announcement of the pandemic, and airline traffic data remained constant until the week of 9 March 2020, when the pandemic was announced (47), and daily Google Trends Mobility data of the countries under study remained stable until 11 March 2020 (48). Therefore, changes in human behaviour and mobility appear to emerge shortly after the pandemic declaration (49,50).

**Figure 1.**
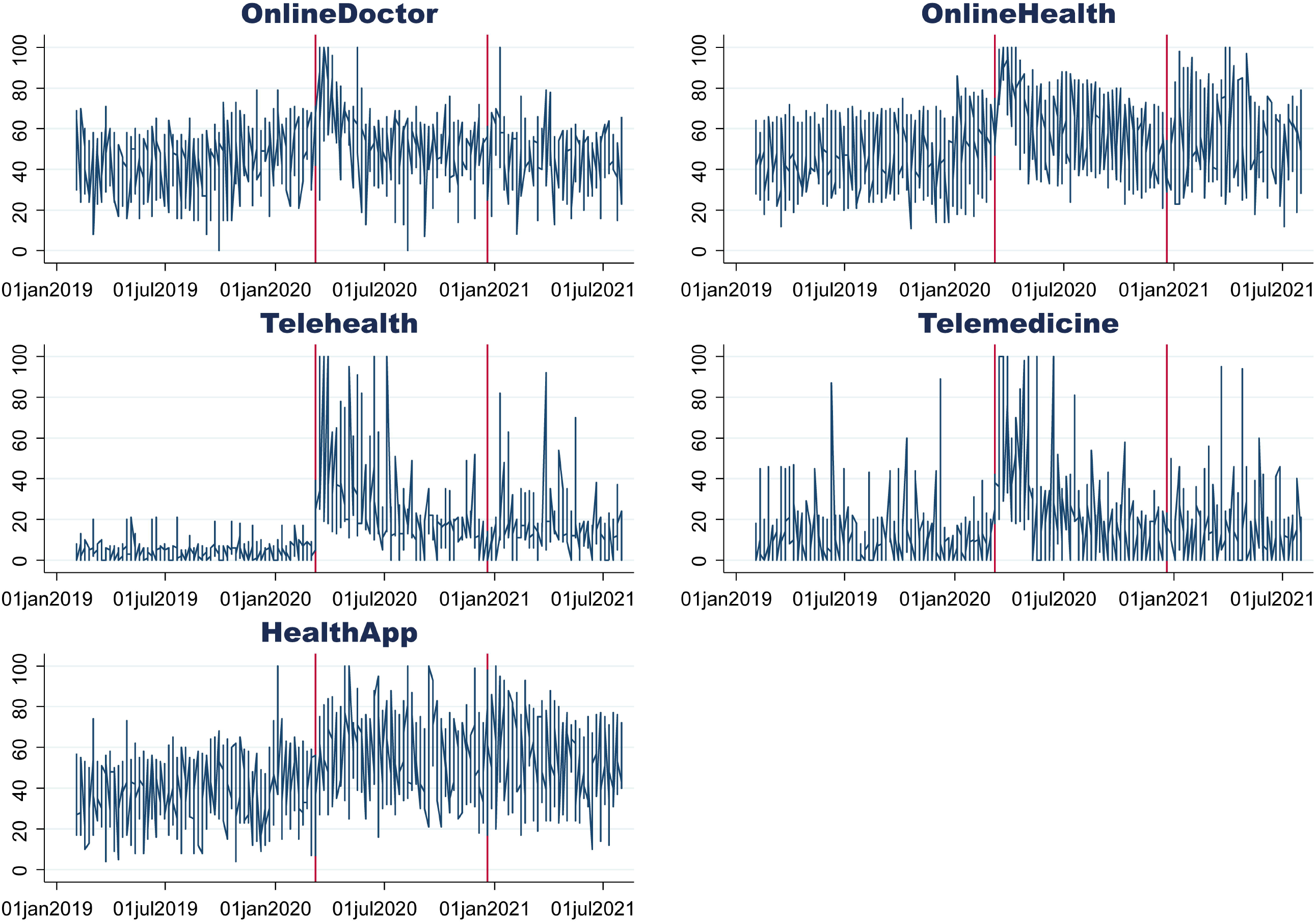
Google Trends search volumes before and after the announcement of the COVID-19 pandemic. The vertical axis shows the average search volume (scaled from 0 to 100) before and after the pandemic was announced and after the announcement of conditional market authorisation of the first COVID-19 vaccines.

### Interrupted time-series analysis

Prior to the announcement of the COVID-19 pandemic, *online doctor* and *online health* showed an increase in relative search volume in Canada, the United Kingdom, and the United States, while *telemedicine* only reported an increasing search volume in the United Kingdom. The search volume of *health app* increased in the United Kingdom and the United States. Upon the announcement of the pandemic, Australia, Canada, the United Kingdom, the United States, and Ireland reported an immediate rise of the search volumes of all five keywords under study. In contrast, New Zealand only reported an immediate rise in *online health, telehealth*, and *telemedicine*. After the immediate shock, Australia and the United States reported a declining trend in the search volumes of all keywords under study except *health app*. Canada reported declining search volumes of *online health, telehealth*, and *telemedicine*, yet an increasing search volume in *health app*. New Zealand also reported declining search volumes of *online health, telehealth*, and *telemedicine*. The United Kingdom showed declining search volumes of *online doctor* and *online health*. Finally, Ireland reported a decline in all keywords under study after the immediate shock of the pandemic announcement. Further details are shown in Figure 2 and Table 1.

**Figure 2.**
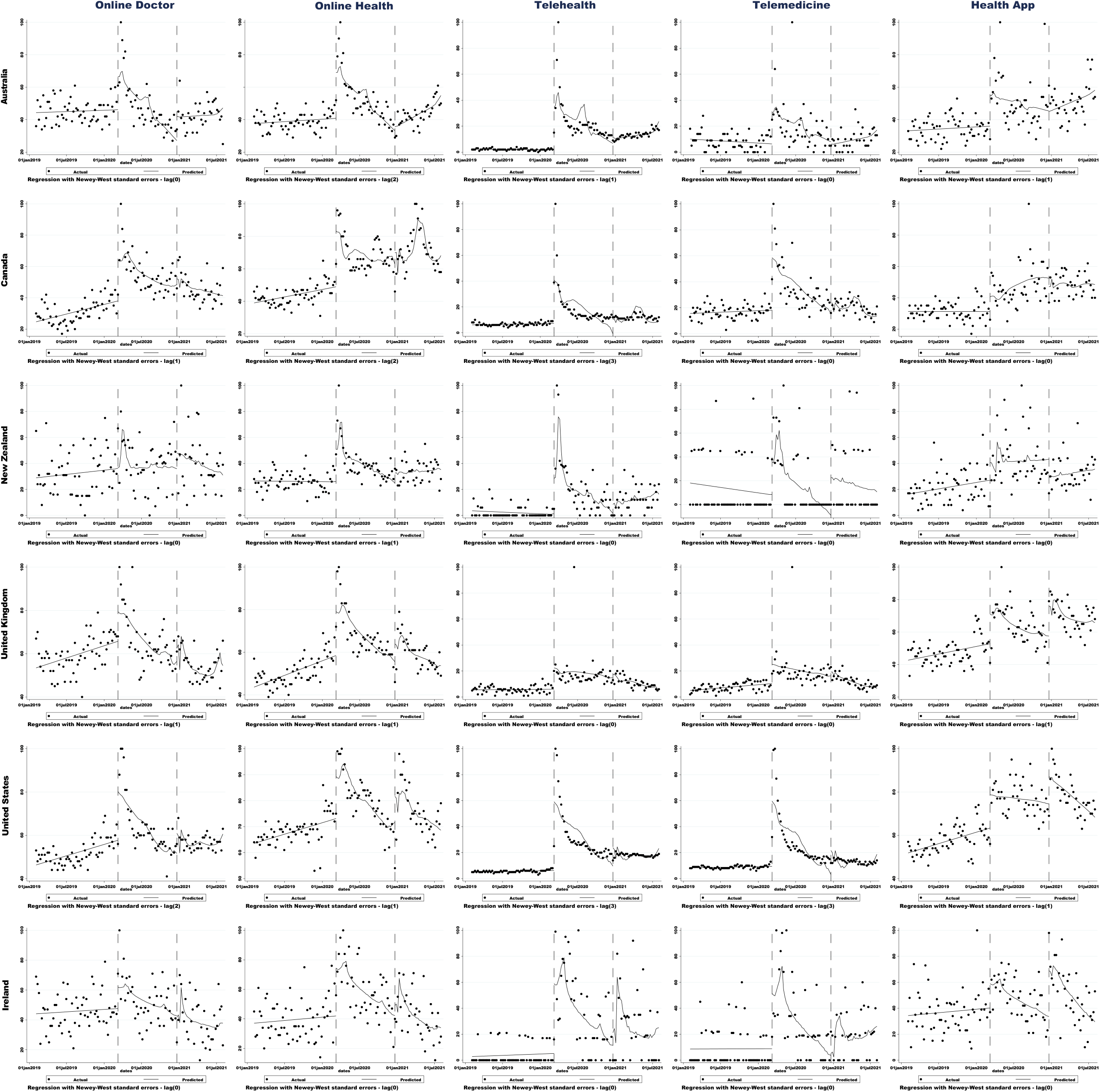
Interrupted time-series regression analysis for the relative search volumes of *online doctor, online health, telehealth, telemedicine*, and *health app* before and after the announcement of the COVID-19 pandemic and the announcement of the first COVID-19 vaccines. The models include seven-day moving averages of reported COVID-19 cases and deaths as covariates. The first interruption occurs at 11 March 2020 and the second at 20 December 2020.

**Table 1.**
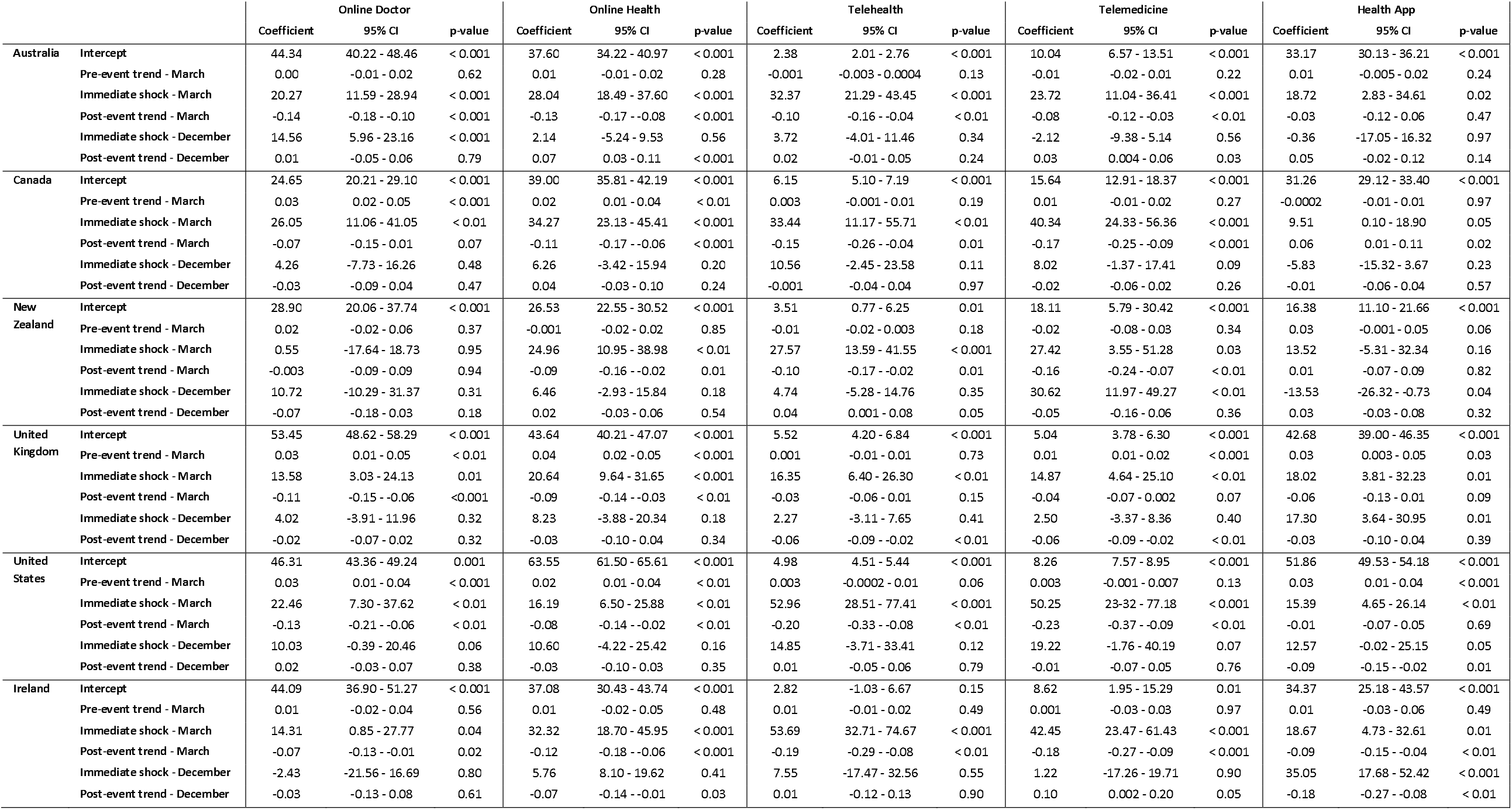
Regression estimates for the relative search volumes of *online doctor, online health, telehealth, telemedicine*, and *health app* before and after the pandemic and the vaccine announcement. The models include seven-day moving averages of reported COVID-19 cases and deaths as covariates.

Following the vaccine announcement on 20 December 2020, the search volume of *telemedicine* in New Zealand reported another immediate rise, while *health app* searches fell. Simultaneously, *health app* search volumes increased in the United Kingdom, the United States, and Ireland. After the immediate impact, Australia reported an upward trend in the search volumes of *online health* and *telemedicine*, while New Zealand reported a positive trend for the search volume of *telehealth*. In the United Kingdom, *telehealth* and *telemedicine* both reported declining trends, as well as *health app* in the United States. Finally, an increasing trend was observed in the search volume of *telemedicine* in Ireland, while *online health* and *health app* decreased. Further details are also shown in Figure 2 and Table 1.

To further check the robustness of our findings, we used data from 2017-2019 and conducted placebo tests with events taking place in the same dates in 2018 (i.e., 08 March 2018, 16 December 2018). eFigure 1 and eTable 1 in Annex A indicate some seasonal shocks in March in *online doctor* and *online health* when using a placebo sample. However, in contrast to our main findings, the shocks in the placebo sample show a downward shock rather than an increase in search volume. While *online health* and *telehealth* reported a positive shock in March in Australia, the main model coefficients are significantly higher than the upper bound of the placebo coefficients’ 95% CI (*online health*, 2.19 - 23.80; *telehealth*, 1.82 - 32.06).

## Discussion

### Summary of key findings

Using Google Trends time-series data, we analysed how digital health search behaviour changed after the pandemic announcement and how it developed during the later stages of the pandemic. We observed an immediate rise in digital health searches in all countries under study, though there is some variability in which keywords were preferred in each country. This is consistent with the claim that healthcare underwent an emergency transition to a digital paradigm when the pandemic started (2). We also observed that – after this immediate spike – digital health search volumes declined again, sometimes towards pre-pandemic levels. The exception to this is the search volume of *health app*, which showed to either remain stable or gradually increase during the pandemic. This can be explained by the notion that all other keywords are meant to substitute traditional healthcare utilisation, while health apps may be used for a wider variety of purposes (e.g., prevention, fitness, managing subclinical health complaints). Additionally, a large volume of health apps was released and advocated for during the pandemic for surveillance purposes (such as recording vaccination status and generating vaccine passports), which may also contribute to explaining the findings of this study. Seeing that vaccines were rolled out in phases across the general population, this can explain why health apps searches remained stable during the entire period under study. Our placebo analysis indicated some statistically significant seasonal trends over the time period 2017-2019. However, these trends were almost exclusively negatively skewed. In the two cases where the trend was positive, our main finding fell outside the confidence interval of the placebo experiment, suggesting that our main results are not driven by an artificial correlation.

### Sustainability of digital health uptake

It is well established that an array of factors need to align in order for a digital innovation to be adopted structurally (2,8), even more so in healthcare which is characteristically one of the slowest domains to adopt digital innovations (51). The beneficial effects of digital health slowly become more apparent and our findings indicate that the general population also seeks out digital healthcare if traditional care is disrupted (2,9). However, our findings indicate that digital healthcare-seeking behaviour is not sustainable at the current stage of progress, seeing how the search volumes under study frequently dropped back to pre-pandemic levels. While this may be partially explained by the return of traditional health services, it also supports previously speculated notions that the digital capacity of the general population may not be sufficiently developed to fully embrace the opportunities brought by digital health (11). Additionally, we highlight how, irrespective of a hypothetical increase in digital competence at a population level over the course of the pandemic, digital skills at later stages in the pandemic continue to be lacking for the purpose of structurally implementing digital health.

This lack of digital readiness extends beyond the general population, as health professionals also continue to voice their concerns regarding the uptake of digital technologies in their practice (7). In light of these reservations, our findings highlight the need for digital (health) training to be developed and implemented in the educational curricula of health professionals. Additionally, training courses for current practitioners may be even more vital, as being informed about the benefits of digital health has been shown to positively influence their decision on using digital health (52). Educational modules and training courses for the general population will also be instrumental in mitigating the effects of the digital health paradox (9).

### Strengths and limitations

This study has a number of strengths. This study is – to our knowledge – the first of its kind that captures the prolonged effect of the COVID-19 pandemic on digital health-seeking behaviour using a comprehensive list of possible search terms, which adequately captures digital health utilisation. While other datasets may exist that capture digital healthcare utilisation, a unique advantage of Google Trends is the high frequency with which it is collected. This allows us to examine both short- and long-term trends from a comparative cross-country perspective. Furthermore, Google Trends data is considered revealed preference data, thus allowing for actual behaviours of Google users to be analysed. Our countries of choice also include a mix of digitally prolific countries (e.g. United Kingdom and Australia) and more digitally hesitant countries (e.g. United States) (9,10), thus providing a balanced representation of search behaviour across countries at various stages of digital health development.

Limitations also need to be considered. While we were not able to ascertain sociodemographic characteristics, we need to consider the possibility of selection bias since young people are more likely to seek health information digitally given their digital skills (11,53). Also, population groups without internet access are unable to contribute to this dataset (11,53). Furthermore, data from only one search engine are included. While Google Trends is the most popular search engine, other search engines may have also been used to seek digital healthcare. Our study does not capture digital health-seeking behaviour of people that directly reached out to their general practitioner or medical contact point. Finally, our data does not allow us to capture the supply-side of digital health delivery (i.e., availability and quality), which may interact with the demand for digital health services.

### Implications for research, policy, practice

Recently, calls for increased and accelerated uptake of digital technologies in healthcare have appeared in various magnitudes (54–57). However, our findings provide a strong signal that such acceleration would be premature and could undermine the real potential of digital health delivery (12). Vulnerable population groups, in particular, may be further disadvantaged and excluded through lack of access to digital infrastructure and underdeveloped digital skills. The recently launched European Health Data Space is an example of how policy frameworks can risk exacerbating existing digital divides if policymakers do not address the underlying lack of access to digital infrastructure and underdeveloped digital skills before structurally rolling out digital health tools (58). As such, capacity-building efforts in the areas of digital infrastructure and skills should be combined with the large-scale introduction of digital health technologies to ensure that citizens and health professionals alike are in the position to recognise the value of digital health tools once they enter the healthcare market at large (7–9,13,52,59). Nevertheless, the deployment of digital health tools should not cease and instead be carefully designed and targeted to specific population groups and health services, because digital health is considered a type of healthcare whose value can only be derived after directly experiencing it (57). Methodically nurturing the readiness and tension for change is, therefore, a vital element in the process of adopting digital health as a mainstream mode of healthcare.

Policy environments are vital to the process of taking up of digital health (9,10). National standards play an important role in creating awareness in markets, setting norms, and safeguarding basic quality dimensions of digital health (10). So long as these factors remain underdeveloped, digital health will face substantial barriers to being implemented and assimilated into national healthcare settings. In fact, the absence of robust market access regulatory frameworks for digital health may also contribute to the current environment where novel tools are stuck in the proof-of-concept or pilot phase and cannot pivot to clinical adoption (3).

Finally, our findings speak to an overarching change in preference for digital health-seeking behaviour. We recognise that the development of digital health is heterogeneous and may differ per health domain. As such, more in-depth analyses of readiness to use digital health are warranted per health domain. Furthermore, educational material should be developed for the purpose of better acquainting the general population with the concept of digital health, building their capacity to harness the potential of digital tools and navigate digital environments (13). More extensive educational material may be prepared for key stakeholder groups, such as patients, health professionals, and policymakers. Closer engagement with the general public and patient groups will further prove to be instrumental in facilitating digital health tools that are widely accessible (9,58).

## Conclusions

This article explores digital health-seeking behaviour in six Anglophone countries at different stages of the COVID-19 pandemic. We show that, after a brief spike, digital health-seeking behaviour frequently declines to levels comparable to before the pandemic; thus, pointing to a lack of readiness to adopt digital health structurally as a mode of healthcare. We highlight potential next steps for digital health integration in the countries under study, while acknowledging the common need for policy innovation, increasing awareness of digital health and its potential, and capacity building (including both digital infrastructure and digital literacy). While the futuristic use of digital technologies in health is promising, we must allow both the policy landscapes and digital health literacy levels to catch up with the rapid advances in technology (2,60). Ultimately, it is evident that more than a pandemic is needed to sustainably implement digital health.

## Supporting information

Annex A

## Data Availability

Data are publicly available from the Google Trends webpage (https://trends.google.com/). The data extraction, cleaning, and analysis code are openly available: https://bit.ly/39ZU0oL

## Acknowledgements

We would like to thank Ms Alicja Mastylak for her time and effort in proofreading this article.

