## Supplementary material for "Has the pandemic enhanced and sustained digital health-seeking behaviour? A big data interrupted time-series analysis of Google Trends": Annex A

eFigure 1. Interrupted time-series regression analysis for the relative search volumes of *online doctor*, *online health*, *telehealth*, *telemedicine*, and *health app* in the time period February 2017 to August 2019. The first interruption occurs at 8 March 2018 and the second at 16 December 2018.

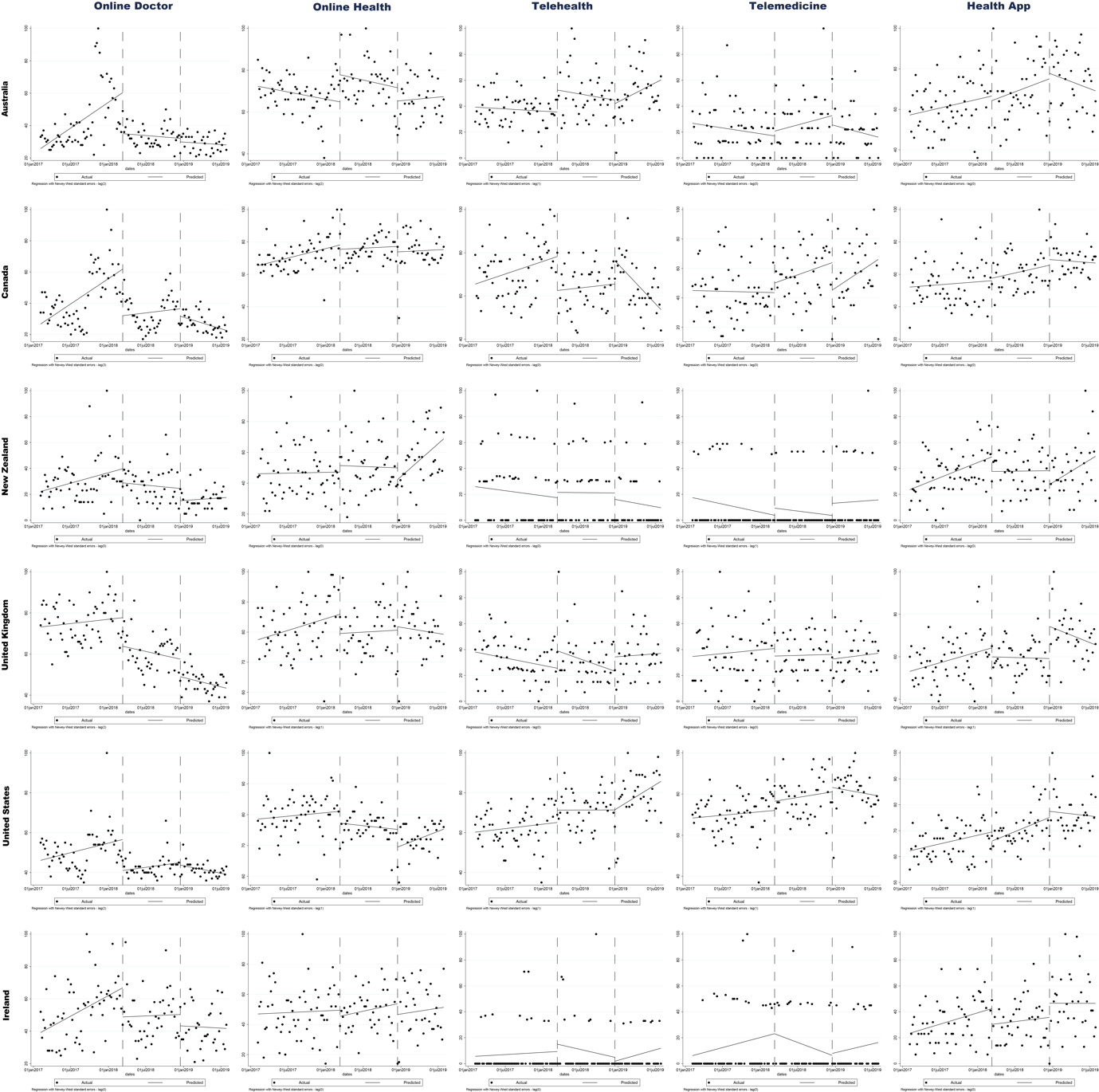

eTable 1.

|  |  | **Online Doctor** | | | **Online Health** | | | **Telehealth** | | | **Telemedicine** | | | **Health App** | | |
| --- | --- | --- | --- | --- | --- | --- | --- | --- | --- | --- | --- | --- | --- | --- | --- | --- |
|  |  | **Coefficient** | **95% CI** | **p-value** | **Coefficient** | **95% CI** | **p-value** | **Coefficient** | **95% CI** | **p-value** | **Coefficient** | **95% CI** | **p-value** | **Coefficient** | **95% CI** | **p-value** |
| **Australia** | **Intercept** | 25.80 | 20.05 - 31.55 | < 0,001 | 72.32 | 68.39 - 76.26 | < 0.001 | 39.25 | 32.72 - 45.78 | < 0.001 | 26.75 | 16.99 - 36.51 | < 0.001 | 57.23 | 51.51 - 62.94 | < 0.001 |
|  | **Pre-event trend - March** | 0,09 | 0,04 - 0,13 | < 0,001 | -0.02 | -0.05 - 0.01 | 0.21 | -0.01 | -0.03 - 0.02 | 0.46 | -0.02 | -0.06 - 0.01 | 0.18 | 0.02 | -0.004 - 0.05 | 0.09 |
|  | **Immediate shock - March** | -25.36 | -40.89 - -9.83 | < 0,001 | 12.99 | 2.19 - 23.80 | 0.02 | 16.94 | 1.82 - 32.06 | 0.03 | 3.66 | -10.19 - 17.53 | 0.60 | -2.28 | -14.91 - 10.35 | 0.72 |
|  | **Post-event trend - March** | -0.01 | -0.04 - 0.02 | 0.50 | -0.02 | -0.06 - 0.02 | 0.30 | -0.03 | -0.10 - 0.04 | 0.43 | 0.04 | -0.04 - 0.13 | 0.33 | 0.04 | -0.03 - 0.11 | 0.26 |
|  | **Immediate shock - December** | -2.16 | -8.15 - 3.84 | 0.48 | -6.36 | -15.76 - 3.04 | 0.18 | -2.74 | -18.29 - 12.81 | 0.73 | -7.19 | -27.41 - 13.02 | 0.48 | 2.70 | -11.34 - 16.74 | 0.70 |
|  | **Post-event trend - December** | -0.01 | -0.03 - 0.01 | 0.42 | 0.01 | -0.06 - 0.07 | 0.78 | 0.08 | -0.02 - 0.18 | 0.11 | -0.04 | -0.13 - 0.05 | 0.36 | -0.04 | -0.11 - 0.03 | 0.28 |
| **Canada** | **Intercept** | 26.66 | 17.27 - 36.05 | < 0,001 | 65.39 | 61.24 - 69.54 | < 0.001 | 65.49 | 59.82 - 71.16 | < 0.001 | 45.01 | 35.83 - 54.19 | < 0.001 | 52.14 | 45.39 - 58.89 | < 0.001 |
|  | **Pre-event trend - March** | 0.09 | 0.04 - 0.13 | < 0,001 | 0.03 | 0.01 - 0.06 | < 0.01 | 0.03 | 0.01 - 0.06 | 0.02 | -0.004 | -0.04 - 0.03 | 0.84 | 0.01 | -0.02 - 0.04 | 0.54 |
|  | **Immediate shock - March** | -29.84 | -47.53 - -12.15 | < 0,01 | -2.67 | -11.89 - 6.56 | 0.57 | -15.72 | -24.95 - -6.49 | < 0.01 | 6.54 | -5.67 - 18.76 | 0.29 | 1.54 | -8.74 - 11.82 | 0.77 |
|  | **Post-event trend - March** | 0.02 | -0,05 - 0.08 | 0.65 | 0.01 | -0.03 - 0.05 | 0.76 | 0.01 | -0.03 - 0.05 | 0.59 | 0.05 | -0.01 - 0.11 | 0.10 | 0.03 | -0.02 - 0.08 | 0.23 |
|  | **Immediate shock - December** | -4.78 | -15.80 - 6.24 | 0.39 | -3.39 | -15.43 - 8.65 | 0.58 | 10.82 | 2.62 - 19.01 | 0.01 | -18.67 | -36.89 - -0.45 | 0.05 | 3.48 | -7.55 - 14.51 | 0.53 |
|  | **Post-event trend - December** | -0.04 | -0.06 - -0.02 | < 0,001 | 0.01 | -0.06 - 0.08 | 0.85 | -0.10 | -0.14 - -0.06 | < 0.001 | 0.09 | -0.04 - 0.22 | 0.16 | -0.01 | -0.06 - 0.04 | 0.72 |
| **New Zealand** | **Intercept** | 22.21 | 16.59 - 27.84 | < 0,001 | 46.13 | 38.11 - 54.16 | < 0.001 | 25.93 | 12.29 - 39.56 | < 0.001 | 17.43 | 7.29 - 27.58 | < 0.01 | 23.62 | 16.03 - 31.20 | < 0.001 |
|  | **Pre-event trend - March** | 0.04 | 0.01 - 0.08 | 0.01 | 0.003 | -0.03 - 0.03 | 0.86 | -0.02 | -0.08 - 0.04 | 0.48 | -0.03 | -0.08 - 0.01 | 0.10 | 0.06 | 0.03 - 0.10 | < 0.001 |
|  | **Immediate shock - March** | -10.91 | -22.85 - 1.03 | 0.07 | 4.20 | -9.45 - 17.86 | 0.54 | 4.12 | -16.79 - 25.04 | 0.70 | 5.51 | -8.21 - 19.22 | 0.43 | -11.31 | -24.13 - 1.52 | 0.08 |
|  | **Post-event trend - March** | -0.02 | -0.06 - 0.03 | 0.53 | -0.01 | -0.07 - 0.06 | 0.87 | -0.002 | -0.09 - 0.09 | 0.97 | -0.02 | -0.08 - 0.04 | 0.51 | 0.002 | -0.06 - 0.06 | 0.94 |
|  | **Immediate shock - December** | -9.43 | -20.46 - 1.59 | 0.09 | -7.58 | -19.90 - 4.73 | 0.23 | -4.91 | -24.38 - 14.57 | 0.62 | 9.32 | -7.22 - 25.86 | 0.27 | -10.68 | -23.79 - 2.43 | 0.11 |
|  | **Post-event trend - December** | 0.01 | -0.03 - 0.05 | 0.59 | 0.12 | 0.04 - 0.19 | < 0.01 | -0.03 | -0.12 - 0.06 | 0.52 | 0.01 | -0.10 - 0.13 | 0.84 | 0.10 | -0.01 - 0.20 | 0.06 |
| **United Kingdom** | **Intercept** | 73.17 | 66.92 - 79.42 | < 0,001 | 77.46 | 73.25 - 81.67 | < 0.001 | 38.10 | 29.24 - 46.96 | < 0.001 | 34.63 | 23.96 - 45.31 | < 0.001 | 53.13 | 48.06 - 58.19 | < 0.001 |
|  | **Pre-event trend - March** | 0.01 | -0.02 - 0.04 | 0.45 | 0.02 | -0.002 - 0.04 | 0.07 | -0.03 | -0.07 - 0.01 | 0.10 | 0.02 | -0.03 - 0.06 | 0.51 | 0.03 | -0.001 - 0.06 | 0.06 |
|  | **Immediate shock - March** | -13.92 | -23.94 - -3.91 | < 0,01 | -6.37 | -13.78 - 1.03 | 0.09 | 13.44 | -3.35 - 30.24 | 0.12 | -5.91 | -20.45 - 8.64 | 0.42 | -4.23 | -13.34 - 4.88 | 0.36 |
|  | **Post-event trend - March** | -0.02 | -0.06 - 0.01 | 0.19 | 0.004 | -0.03 - 0.03 | 0.80 | -0.06 | -0.14 - 0.03 | 0.18 | 0.01 | -0.06 - 0.07 | 0.88 | -0.003 | -0.03 - 0.03 | 0.86 |
|  | **Immediate shock - December** | -8.54 | -14.31 - -2.77 | < 0,01 | 1.14 | -9.88 - 12.16 | 0.84 | 11.02 | -5.87 - 27.92 | 0.20 | -3.60 | -17.65 - 10.45 | 0.61 | 15.11 | 3.14 - 27.07 | 0.01 |
|  | **Post-event trend - December** | -0.02 | -0.05 - 0.002 | 0.07 | -0.01 | -0.08 - 0.05 | 0.73 | 0.01 | -0.08 - 0.10 | 0.79 | 0.02 | -0.05 - 0.09 | 0.59 | -0.04 | -0.11 - 0.03 | 0.28 |
| **United States** | **Intercept** | 46.13 | 40.72 - 51.54 | < 0,001 | 78.58 | 74.39 - 82.76 | < 0.001 | 60.27 | 55.14 - 65.39 | < 0.001 | 68.36 | 63.63 - 73.09 | < 0.001 | 62.24 | 59.27 - 65.20 | < 0.001 |
|  | **Pre-event trend - March** | 0.03 | -0.002 - 0.05 | 0.07 | 0.01 | -0.01 - 0.02 | 0.50 | 0.01 | -0.02 - 0.04 | 0.42 | 0.01 | -0.02 - 0.03 | 0.47 | 0.02 | 0.0002 - 0.04 | 0.05 |
|  | **Immediate shock - March** | -15.57 | -23.72 - -7.42 | < 0,001 | -3.84 | -8.94 - 1.26 | 0.14 | 6.22 | -4.14 - 16.59 | 0.24 | 4.49 | -4.20 - 13.18 | 0.31 | -3.53 | -9.48 - 2.41 | 0.24 |
|  | **Post-event trend - March** | 0.01 | -0.01 - 0.04 | 0.22 | -0.01 | -0.03 - 0.01 | 0.49 | 0.0003 | -0.04 - 0.05 | 0.99 | 0.02 | -0.02 - 0.05 | 0.38 | 0.03 | 0.01 - 0.05 | < 0.01 |
|  | **Immediate shock - December** | -1.23 | -6.68 - 4.22 | 0.66 | -5.72 | -11.49 - 0.05 | 0.05 | 0.10 | -14.31 - 14.50 | 0.99 | 2.16 | -8.79 - 13.10 | 0.70 | 2.36 | -6.46 - 11.19 | 0.60 |
|  | **Post-event trend - December** | -0.02 | -0.04 - -0.002 | 0.03 | 0.03 | -0.01 - 0.06 | 0.10 | 0.06 | -0.02 - 0.15 | 0.14 | -0.02 | -0.08 - 0.04 | 0.55 | -0.01 | -0.06 - 0.05 | 0.75 |
| **Ireland** | **Intercept** | 39.22 | 31.73 - 46.72 | < 0,001 | 46.97 | 37.63 - 56.31 | < 0.001 | 5.64 | -1.36 - 12.63 | 0.11 | 6.19 | -3.00 - 15.38 | 0.19 | 23.47 | 16.46 - 30.49 | < 0.001 |
|  | **Pre-event trend - March** | 0.07 | 0.04 - 0.10 | < 0,001 | 0.01 | -0.03 - 0.05 | 0.76 | 0.01 | -0.02 - 0.04 | 0.58 | 0.04 | -0.01 - 0.09 | 0.08 | 0.05 | 0.01 - 0.08 | 0.01 |
|  | **Immediate shock - March** | -18.01 | -31.32 - -4.69 | < 0,01 | -4.03 | -15.46 - 7.40 | 0.49 | 5.69 | -11.82 - 23.19 | 0.52 | -0.07 | -20.92 - 20.78 | 0.99 | -12.19 | -24.87 - 0.49 | 0.06 |
|  | **Post-event trend - March** | 0.01 | -0.06 - 0.07 | 0.87 | 0.03 | -0.02 - 0.08 | 0.22 | 0.04 | -0.13 - 0.05 | 0.42 | -0.06 | -0.15 - 0.03 | 0.20 | 0.02 | -0.04 - 0.08 | 0.51 |
|  | **Immediate shock - December** | -7.00 | -19.79 - 5.78 | 0.28 | -7.16 | -23.78 - 9.46 | 0.40 | -3.01 | -18.29 - 12.27 | 0.70 | 1.50 | -17.08 - 20.07 | 0.87 | 10.93 | -8.09 - 29.95 | 0.26 |
|  | **Post-event trend - December** | -0.01 | -0.07 - 0.06 | 0.83 | 0.02 | -0.08 - 0.12 | 0.68 | 0.04 | -0.02 - 0.11 | 0.17 | 0.04 | -0.06 - 0.13 | 0.42 | -0.001 | -0.11 - 0.11 | 0.99 |
